# Comparison of indicators for assessing wasting among children younger than 5 years: a longitudinal study in northern Kenya

**DOI:** 10.1101/2024.08.14.24311972

**Authors:** Calistus Wilunda, Faith Thuita, Bonventure Mwangi, Valerie L. Flax, Chessa K. Lutter, Linda Adair, Dickson Amugsi, Hazel Odhiambo, Esther Anono, Albert Webale, Gillian Chepkwony, Stephen Ekiru, Elizabeth Kimani-Murage, Estelle Sidze

## Abstract

Mid-upper arm circumference (MUAC) or weight-for-height/length Z-score (WHZ) are recommended in wasting diagnosis, but there are discrepancies between these indicators in identifying children as wasted. We compared the extent to which WHZ, MUAC, MUAC-for-age Z-score (MAZ) identify the same children as wasted and assessed the predictors of discordance and concordance in wasting diagnosis by these indicators using data from a longitudinal study of children younger than 3 years at recruitment in Turkana and Samburu counties. Wasting prevalence was consistently lower based on MUAC than WHZ and MAZ. Compared to WHZ, MAZ had higher sensitivity than MUAC, with the sensitivity of MAZ increasing and MUAC decreasing with age. Both indicators had high specificity. WHZ had a better agreement with MAZ than MUAC in wasting diagnosis. Older children were less likely to be classified as wasted by MUAC alone or by both MUAC and WHZ but were more likely to be classified as wasted by WHZ alone, MAZ alone or by both MAZ and WHZ. Compared to girls, boys were less likely to be classified as wasted by MUAC alone but more likely to be classified as wasted by WHZ alone. Stunted children were more likely to be classified as wasted by MUAC alone, MAZ alone, both MUAC and WHZ, and both MAZ and WHZ but not by WHZ alone. Classifications of wasting based on WHZ, MAZ, and MUAC are age, sex, and stunting status dependent. Compared to WHZ, MAZ is a more reliable and valid indicator than MUAC in these settings.

## INTRODUCTION

Child wasting (low weight-for-height) is a persistent public health challenge. Globally, approximately 45 million children under 5 were affected by wasting in 2022 (WHO, 2023), with more than 97% of them living in Asia and Africa (WHO, 2023). Wasting is the most dangerous form of undernutrition in children in the short term as it has a stronger association with morbidity and mortality compared to stunting and underweight (Olofin et al., 2013). Because there is no single gold-standard for assessing wasting, the World Health Organization (WHO) recommends the use of mid-upper arm circumference (MUAC) or weight-for-height/length Z-score (WHZ) to identify wasted children for admission into and discharge from nutritional rehabilitation programs (WHO, 2023). MUAC is also used to monitor the prevalence of wasting in surveillance programs. MUAC has operational advantages compared to WHZ including not requiring a reference table, not involving cumbersome and expensive equipment, requiring only one measurement, and ease of use and interpretation by community health workers and families with limited education – facilitating its use in community-based programs and humanitarian settings.

However, several studies have shown discrepancies between WHZ and MUAC in identifying wasted children (Bilukha & Leidman, 2018; Leidman, Couture, Hulland, & Bilukha, 2019; Roberfroid et al., 2015). A large-scale pooled analysis of data from cross-sectional surveys found that WHZ and MUAC measurements identify different sets of children with wasting (Custodio et al., 2022). The analysis also showed that wasting prevalence was lower when using MUAC than when using WHZ. Child’s sex, age, and stunting status are some of the factors that influence whether children are diagnosed as acutely malnourished by the different indicators (Custodio et al., 2018; Custodio et al., 2022). While use of a single MUAC cut point of 12.5 cm has been used for simplicity to identify wasted children, it does not take age and sex differences into account, leading to potential misclassification related to these variables relative to WHZ. Thus, the use of MUAC adjusted for age [(MUAC-for-age Z-score (MAZ)] has been suggested as a more useful indicator of wasting status. An analysis of cross-sectional data from Somalia showed that MAZ yielded wasting prevalence estimates similar to those obtained by WHZ (Custodio et al., 2018).

Almost all studies comparing different anthropometric indicators of wasting are cross-sectional and there is a dearth of information on how the performance of the three indicators change over time in a cohort of children. Moreover, few studies have directly compared the performance of MAZ to WHZ in identifying wasted children aged 0 to 59 months. Using data from a longitudinal study, we aimed to compare the extent to which MUAC, MAZ and WHZ identify the same children as wasted and to assess the predictors of discordance and concordance of WHZ with MUAC and MAZ in identifying wasted children.

**KEY MESSAGES**

- Wasting prevalence was consistently lower when measured using MUAC than WHZ and MAZ.
- With reference to WHZ, MAZ was a more valid and reliable indicator in identifying wasted children than MUAC.
- Classifications of wasting based on WHZ, MAZ, and MUAC are age, sex, and stunting status dependent.

## METHODS

This study used data from the Nawiri Longitudinal Study, a 24-month mixed-methods observational study in Turkana and Samburu counties in northern Kenya. Both counties are classified as arid and semiarid lands (ASALs) and are characterized by a hot and dry climate, unreliable rainfall patterns, and frequent droughts. Details of this study are described elsewhere (Wilunda et al., (Under review)). In brief, children younger than 3 years and their mothers/caregivers were recruited and followed approximately every 4 months for six waves of data collection as shown in **Supplemental Tables 1 and 2**. The timing of data collection was meant to capture seasonality, however, there was a prolonged drought after Wave 1 in both counties and the drought situation at Waves 2-5 was at “alarming” or “emergency” levels, while that at Wave 6, it was at the “alerting” level in most livelihood zones in the two counties according to the National Drought Management Authority (**Supplemental Tables 1 and 2)**. Anthropometric measurements (weight, length/height, and MUAC) of all children under 5 years in the sampled households were taken at Wave 1, and one child under 3 years was randomly selected and followed at Waves 2 to 6.

A sample size of 1,544 and 669 households in Turkana and Samburu, respectively, was calculated to provide wasting estimates at the county level, as previously described (Wilunda et al., (Under review)). The number of households was allocated proportionally to the population size of each stratum (survey zone). Turkana has four survey zones (Central, North, West, and South) while Samburu has three (North, Central, and East).

A representative sample of children under 3 years and their mothers/caregivers was obtained using multistage sampling, stratified by survey zone. Villages were treated as clusters within a survey zone, from which a random sample of 25 and 20 villages in Turkana and Samburu, respectively, was drawn. Simple random sampling was then used to select study households following a household listing to identify eligible households (those with children under 3 years old).

Data were collected between May 2021 and September 2023 by experienced and trained fieldworkers using a questionnaire loaded in SurveyCTO. Details of the quality control measures employed in the study have been described elsewhere (Wilunda et al., (Under review)).

Anthropometric data were collected according to recommendations by United Nations Children’s Fund (UNICEF) and WHO (WHO & UNICEF, 2019). The weight of children was measured using a calibrated digital electronic mother/caregiver–child pair weighing scale (Seca 874–200kg). For very young children or those who could not stand on the weighing scale, weight was measured using tared weighing, whereby the weight of the mother or caregiver was measured first, after which she was asked to stand on the scale holding the child. The child’s weight was obtained by subtracting the mother’s or caregiver’s weight from the combined weight of the mother/caregiver and child. Child recumbent length (<2 years) or standing height (≥2 years) was measured using length board (wooden: length/height to 130 cm). The measuring boards were calibrated using piping of a known length, while each scale was tested with a standard weight of 5 kg. Each of the weight and height measurements was taken twice and an average was calculated to ensure accuracy. Child’s MUAC was measured to the nearest 0.1 cm using UNICEF-simplified MUAC tapes (UNICEF, 2019).

Wasting was defined using three indicators: 1) WHZ < –2 standard deviations [SD], 2) MUAC < 12.5 centimeters (cm), and 3) MAZ (< –2 SD) (WHO Multicentre Growth Reference Study Group, 2006). We created three outcome variables: a continuous variable of the difference between WHZ and MAZ, a polychotomous variable of wasting status based on WHZ and MUAC (not wasted based on both WHZ and MUAC, wasted based on MUAC only, wasted based on WHZ only and wasted based on both MUAC and WHZ), and a polychotomous variable of wasting status based on WHZ and MAZ (not wasted based on both WHZ and MAZ, wasted based on MAZ only, wasted based on WHZ only and wasted based on both MAZ and WHZ). Exposure variables in the assessment of the discordance and concordance between different anthropometric indicators in wasting diagnosis included child’s age (months), child’s sex, livelihood zone (pastoral, agro-pastoral, fisher folk (only in Turkana), and urban/peri-urban) and stunting (height-for-age z-score [HAZ] < –2 SD (WHO Multicentre Growth Reference Study Group, 2006). These variables are known to influence the discordance and concordance of different anthropometric indicators in wasting diagnosis (Custodio et al., 2018; Custodio et al., 2022; Leidman et al., 2019; Zaba, Nyawo, & Álvarez Morán, 2020). Other variables included survey wave (Waves 1-6) and survey zone (Central, North, South, and West in Turkana and Central, North and East in Samburu).

### Statistical analysis

Analysis was conducted by county. We summarized children’s characteristics using descriptive statistics. We present the mean ± SD of MUAC, MAZ and WHZ and the prevalence of wasting, based on these indicators, by child’s sex and survey wave. We assessed the validity of MUAC and MAZ in identifying wasted children using sensitivity and specificity, with reference to WHZ. We also calculated the positive predictive values (PPV) and negative predictive values (NPV) of MUAC and MAZ in diagnosing wasting using WHZ as the reference. We assessed the reliability (repeatability) of MUAC and MAZ with reference to WHZ using Pearson’s correlation coefficients. We assessed the level of agreement between WHZ and each of MUAC and MAZ in identifying wasted children using the Kappa statistic, which is a measure of agreement between two tests that is beyond chance alone. Kappa values of <0.4 denoted poor agreement (Celentano & Szklo, 2019). We calculated the difference between WHZ and MAZ (WHZ-MAZ), summarized this variable using the mean ± SD and assessed its predictors using multivariable multilevel linear regression models. We assessed factors associated with concordant and discordant classification of wasting by WHZ vs. MUAC and WHZ vs. MAZ using multivariable multinomial logistic regression. No wasting based on both WHZ and MUAC and both WHZ and MAZ was treated as the base category in the respective models. The effects represent the relative risk of being in the specified group compared to the base group. All multivariable models included survey wave, child’s age, child’s sex, stunting status, survey zone and livelihood zone, and were clustered at the level of the child to account for repeated measures. All descriptive statistics (means and percentages) were weighted to account for the sampling design. Analyses were performed in Stata 19.

### Ethics statement

The African Population and Health Research Center (APHRC) obtained ethical and research approvals and research permits from Amref Health Africa’s Ethical and Scientific Review Committee (Amref ESRC P905/2020) and the National Commission for Science, Technology, and Innovation of Kenya and signed a reliance agreement with RTI International’s Institutional Review Board for the research.

## RESULTS

1,211 and 586 children in Turkana and Samburu, respectively, were recruited in the study at Wave 1. Of these, the percentage surveyed from Waves 2 to 6 was 88.2%, 84.1%, 93.6%, 89.3% and 95.5% in Turkana and 94.9%, 89.1%, 93.7%, 92.7%, and 92.8% in Samburu. Selected characteristics of participants by county and survey wave are summarized in **Supplemental Table 3**. At recruitment, the mean ± SD age of children was 16.8 ± 10.6 months and 14.4 ± 10.0 months in Turkana and Samburu, respectively. Slightly less than half were girls. The prevalence of stunting ranged from 25.9% at Wave 1 to 40.4% at Wave 4 in Turkana and from 24.4% at Wave 1 to 43.6% at Wave 5 in Samburu. In both Turkana and Samburu, the mean WHZ and MAZ generally decreased between Waves 1 and 5 followed by a slight increase at Wave 6 (**Supplemental Table 4**). In contrast, in both counties, the mean MUAC increased between Waves 1 and 6. The mean WHZ and MAZ were generally lower in boys than girls while the mean MUAC was generally higher in boys than girls irrespective of county.

**Figure 1** shows the prevalence of wasting based on WHZ, MUAC, and MAZ by county and survey wave. In Turkana, both WHZ and MAZ indicators showed a higher wasting prevalence than MUAC. Based on WHZ, wasting prevalence was similar at Wave 1 (21.8%) and Wave 6 (22.1%), having peaked at Wave 3 (28.4%). MAZ results showed a similar trend but with a peak at Wave 5 (31.2%). In contrast, MUAC results showed decreasing wasting prevalence between Waves 1 and 6 (13.8 % to 2.5 %). In Samburu, the prevalence of wasting was highest when measured by WHZ, followed by MAZ and MUAC across survey waves.

**Figure 1:**
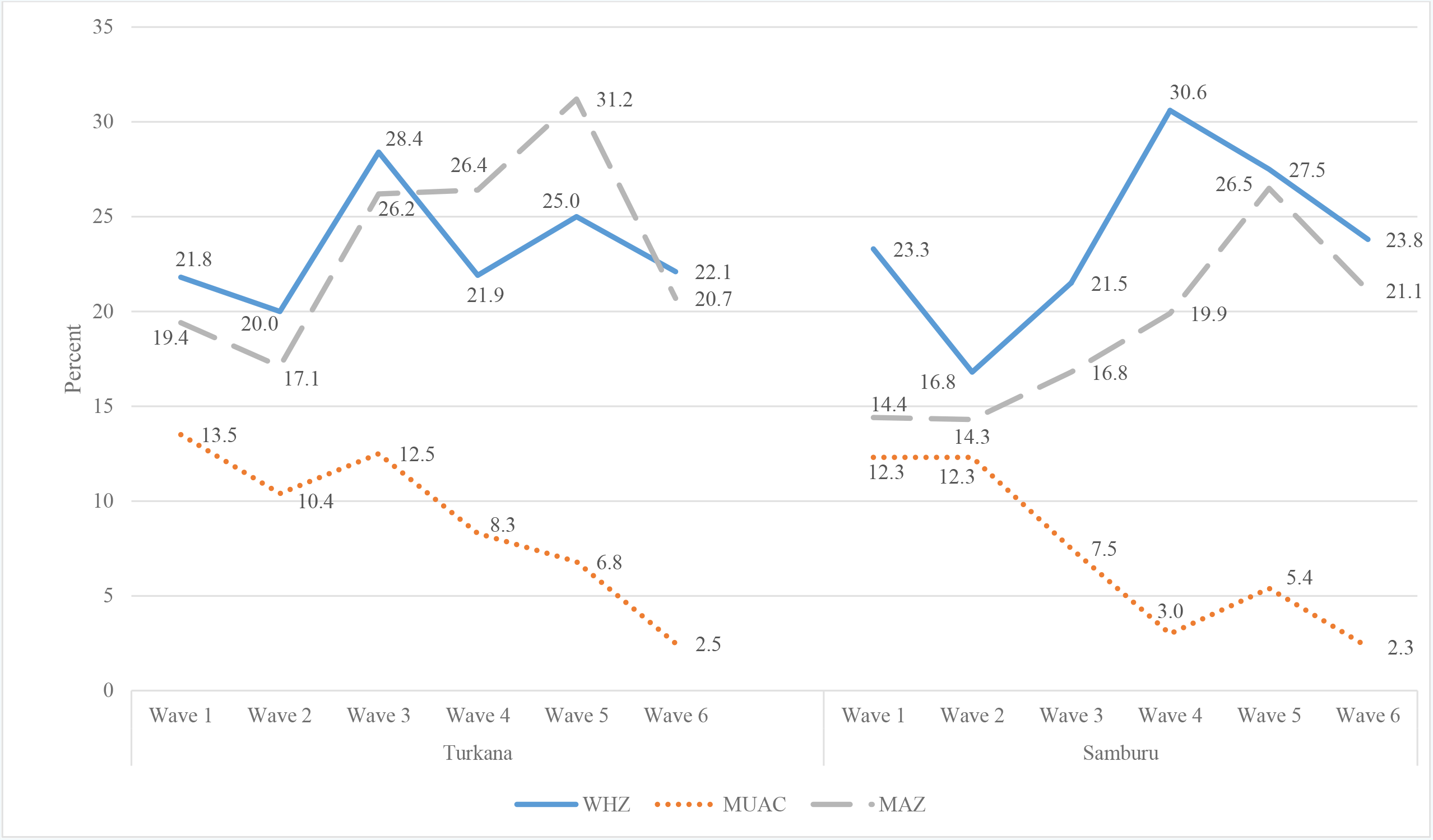
Prevalence of wasting based on WHZ, MUAC and MAZ by county

Based on WHZ, the prevalence of wasting decreased from 23.3% at Wave 1 to 16.8% at Wave 2 followed by an increase to 30.6% at Wave 4 before decreasing slightly to 23.8% at Wave 6. The prevalence based on MAZ generally followed a similar pattern. In contrast, when measured by MUAC, the prevalence of wasting decreased from 12.3% at Wave 1 to 2.3% at Wave 6. There was a time lagged effect of rainfall on wasting prevalence in Turkana: higher rainfall was followed by higher wasting prevalence at the subsequent wave and vice versa (**Supplemental Figure 1**). No clear seasonal pattern was observed in Samburu, however, the lowest rainfall amount in September 2022 (Wave 4) and February/March 2023 (Wave 5) coincided with the highest wasting prevalence at these time points.

The prevalence of wasting based on WHZ and MAZ was generally higher in males than females, but the results based on MUAC varied (**Supplemental Table 5**). In all survey waves, WHZ showed a similar correlation with MAZ and MUAC in both counties (**Supplemental Table 6**). Relative to WHZ, Pearson’s correlation coefficients ranged from 0.472 to 0.735 for MUAC and 0.496 to 0.797 for MAZ.

An assessment of the mean of the difference between WHZ and MAZ showed variation by wave and age category within wave (**Supplemental Table 7**). The mean difference was greater among children aged >24 months than among those aged ≤ 24 months.

**Table 1** and **Supplemental Figures 2 and 3** show the results of sensitivity, specificity and predictive values of MUAC and MAZ with reference to WHZ in the diagnosis of wasting. Both MUAC and MAZ had a similar sensitivity at Wave 1 but diverged thereafter. In Turkana, the sensitivity of MAZ increased from 38.7% at Wave 1 to 60.2% at Wave 4 before decreasing to 50.9% at Wave 6. In contrast, the sensitivity of MUAC was generally low and gradually decreased from 34.5% at Wave 1 to 9.1% at Wave 6 **(Table 2 and Supplemental Figure 2)**. Across survey waves, the specificity of MUAC remained above 91% while that of MAZ remained above 84%. The PPV of MUAC and MAZ generally increased between Waves 1 and 6, albeit with fluctuations in between. Across all waves, MUAC had higher PPV than MAZ. The NPV of MUAC ranged from 77.5% to 82.7% while that of MAZ ranged from 81.0% to 86.3%. In Samburu, between Waves 1 and 6, the sensitivity of MUAC decreased (33.3% to 7.8%) while that of MAZ increased (38.3% to 56.3%), peaking at Wave 5 (**Table 1** and **Supplemental Figure 3**). The specificity of both MUAC and MAZ remained above 91%. The NPV of MAZ remained above 88% while that of MUAC ranged from 72.8% to 90.7%. The PPV of MUAC ranged from 51.0% and 66.7% while that of MAZ ranged from 51.8% to 65.1%.

**Table 1:** Sensitivity, specificity, predictive values of MUAC and MAZ with reference to WHZ in wasting diagnosis.

|  | MUAC |  |  |  |  | MAZ |  |  |  |  |
| --- | --- | --- | --- | --- | --- | --- | --- | --- | --- | --- |
|  | N | Sensitivity (%) | Specificity (%) | PPV (%) | NPV (%) | N | Sensitivity (%) | Specificity (%) | PPV (%) | NPV (%) |
| **Turkana** |  |  |  |  |  |  |  |  |  |  |
| Wave 1 | 913 | 34.5 | 91.8 | 58.5 | 80.7 | 884 | 38.7 | 89.2 | 55.1 | 81.0 |
| Wave 2 | 1,007 | 32.9 | 95.2 | 67.3 | 82.7 | 1,006 | 45.5 | 92.0 | 62.9 | 85.0 |
| Wave 3 | 943 | 30.9 | 96.7 | 79.0 | 77.8 | 938 | 56.6 | 89.3 | 67.7 | 83.8 |
| Wave 4 | 1,079 | 24.8 | 97.6 | 75.9 | 80.8 | 1,065 | 58.7 | 86.3 | 57.1 | 87.1 |
| Wave 5 | 1,035 | 17.9 | 99.0 | 85.7 | 77.5 | 1,035 | 60.4 | 84.9 | 58.3 | 86.0 |
| Wave 6 | 1,026 | 9.1 | 99.0 | 72.4 | 78.9 | 1,006 | 50.9 | 91.5 | 63.9 | 86.3 |
| **Samburu** |  |  |  |  |  |  |  |  |  |  |
| Wave 1 | 434 | 33.3 | 94.9 | 51.3 | 89.9 | 434 | 38.3 | 95.5 | 57.5 | 90.6 |
| Wave 2 | 493 | 38.8 | 94.1 | 51.0 | 90.7 | 491 | 43.3 | 94.6 | 55.8 | 91.3 |
| Wave 3 | 478 | 22.5 | 96.3 | 51.6 | 87.7 | 478 | 40.8 | 93.4 | 51.8 | 90.0 |
| Wave 4 | 545 | 14.2 | 98.4 | 68.2 | 72.8 | 545 | 50.9 | 93.4 | 65.1 | 88.7 |
| Wave 5 | 543 | 16.8 | 98.6 | 73.9 | 83.8 | 543 | 64.4 | 91.6 | 63.7 | 91.8 |
| Wave 6 | 529 | 7.8 | 99.1 | 66.7 | 81.6 | 529 | 56.3 | 92.0 | 63.0 | 89.7 |

**Table 2:**
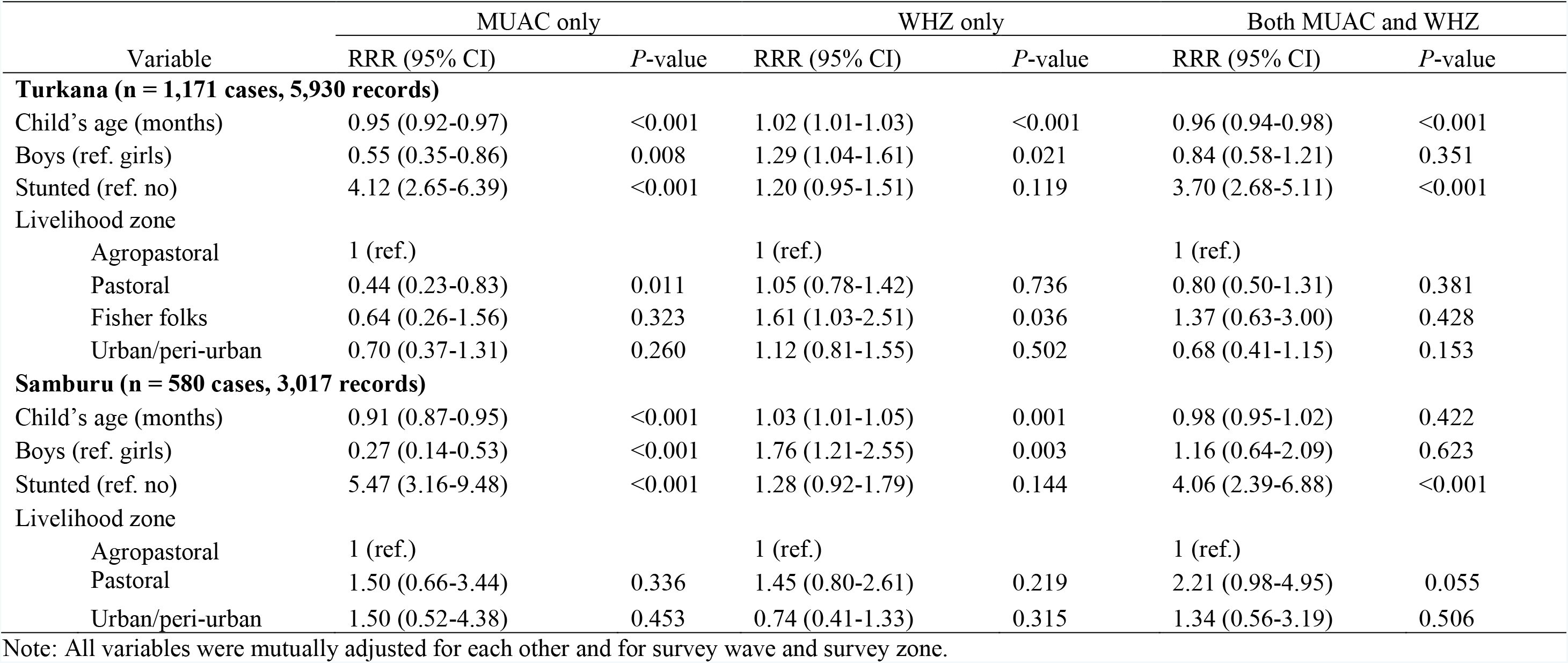
Factors associated with discordance and concordance between MUAC and WHZ in wasting diagnosis.

In both counties, there was poor agreement between WHZ and MUAC (kappa <0.4) across all survey waves except at Wave 1 in Samburu (**Figure 2**). The agreement between WHZ and MUAC generally decreased between Waves 1 and 6. The agreement between WHZ and MAZ was generally good (kappa ≥ 0.4) across all waves except Wave 1 in Turkana and Wave 3 in Samburu.

**Figure 2:**
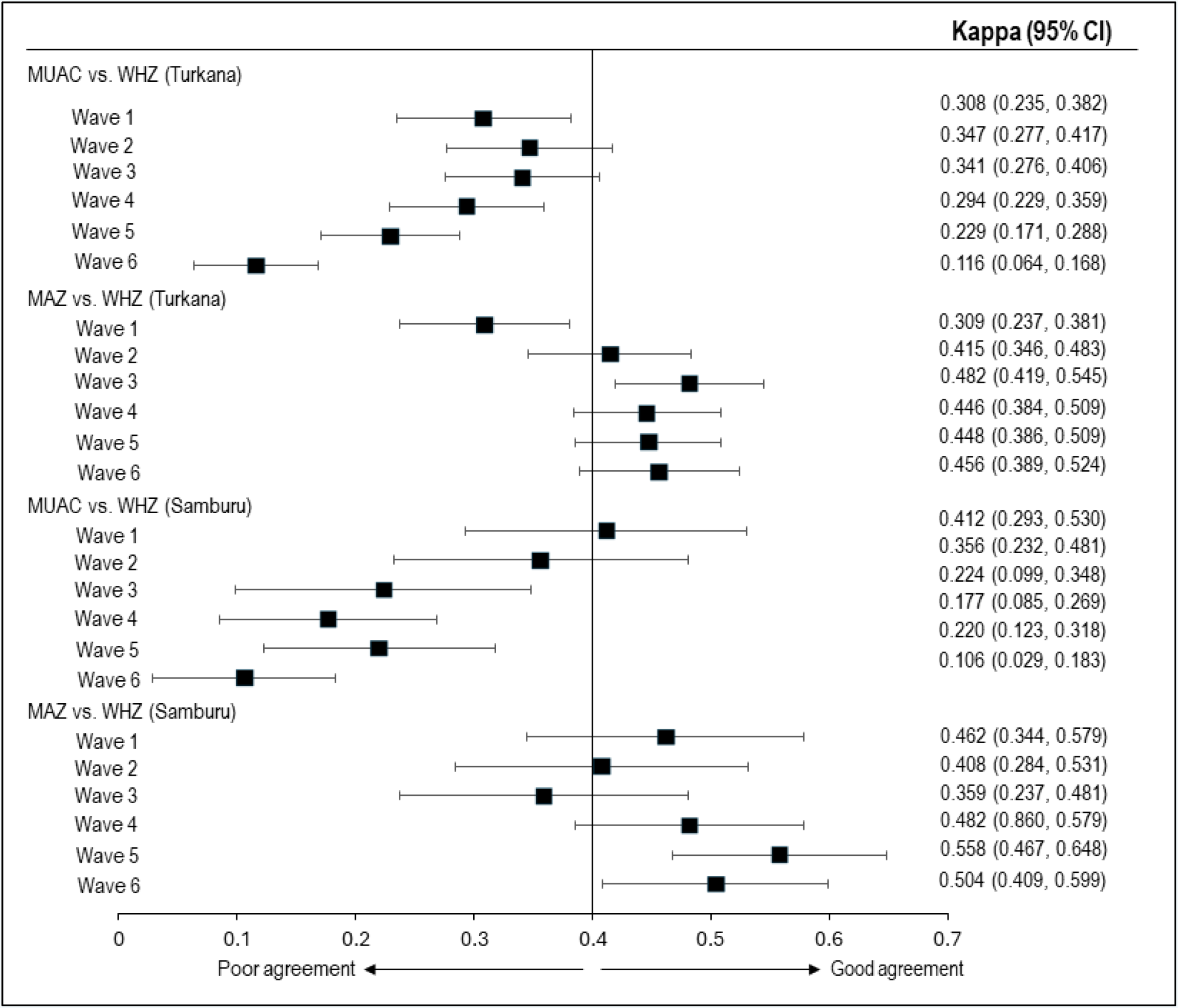
Agreement of MUAC and MAZ with WHZ in identifying children with wasting

The percentage of children classified as wasted based on a combination of different anthropometric indicators is shown in **Supplemental Table 8**. The highest percentage of wasted children were identified by WHZ only, followed by both WHZ and MAZ, and MAZ only. MUAC or MUAC and WHZ together identified fewer wasted children.

**Table 2** shows the determinants of the discordance and concordance between MUAC and WHZ in identifying wasted children. On average, across all surveys, a unit increase in child’s age (months) was associated with 5% and 9% reduced probability of wasting diagnosis by MUAC only in Turkana and Samburu, respectively. The probability of wasting diagnosis by MUAC only was 45% and 73% lower in boys than girls in Turkana and Samburu, respectively and 312% and 447% higher in stunted children than non-stunted children in Turkana and Samburu, respectively. A unit increase in child’s age was associated with 2% and 3% increased likelihood of wasting diagnosis by WHZ only in Turkana and Samburu, respectively. Compared to girls, boys were 29% and 76% more likely to be diagnosed as wasted by WHZ alone in Turkana and Samburu, respectively. Stunting was not associated with wasting diagnosis by WHZ alone. However, it was associated with about a 4-fold increased likelihood of wasting diagnosis by both WHZ and MUAC. Older children were less likely to be diagnosed as wasted by both WHZ and MUAC in Turkana. There was no association between child’s sex and wasting diagnosis by both WHZ and MUAC in both counties.

**Table 3** shows the determinants of the discordance and concordance between MAZ and WHZ in identifying wasted children. A unit increase in child’s age was associated with increased likelihood of wasting diagnosis by MAZ alone and both MAZ and WHZ but not by WHZ alone in both counties. Stunting was associated with increased likelihood of wasting diagnosis by MAZ alone and both MAZ and WHZ in both Turkana and Samburu. Compared to girls, boys were 29% and 55% more likely to be diagnosed as wasted by MAZ alone in Turkana and Samburu, respectively, and 87% more likely to be diagnosed as wasted by both MAZ and WHZ in Samburu. Child’s age and stunting were associated with significantly higher mean differences between WHZ and MAZ (WHZ minus WAZ) (**Table 3**). A unit increase in child’s age was associated with an increase of 0.07 and 0.01 difference between WHZ and MAZ in Turkana and Samburu, respectively. Compared to non-stunted children, stunted children had on average 0.40 and 0.32 higher WHZ and MAZ difference in Turkana and Samburu, respectively. The difference between WHZ and WAZ was not influenced by child’s sex.

**Table 3:**
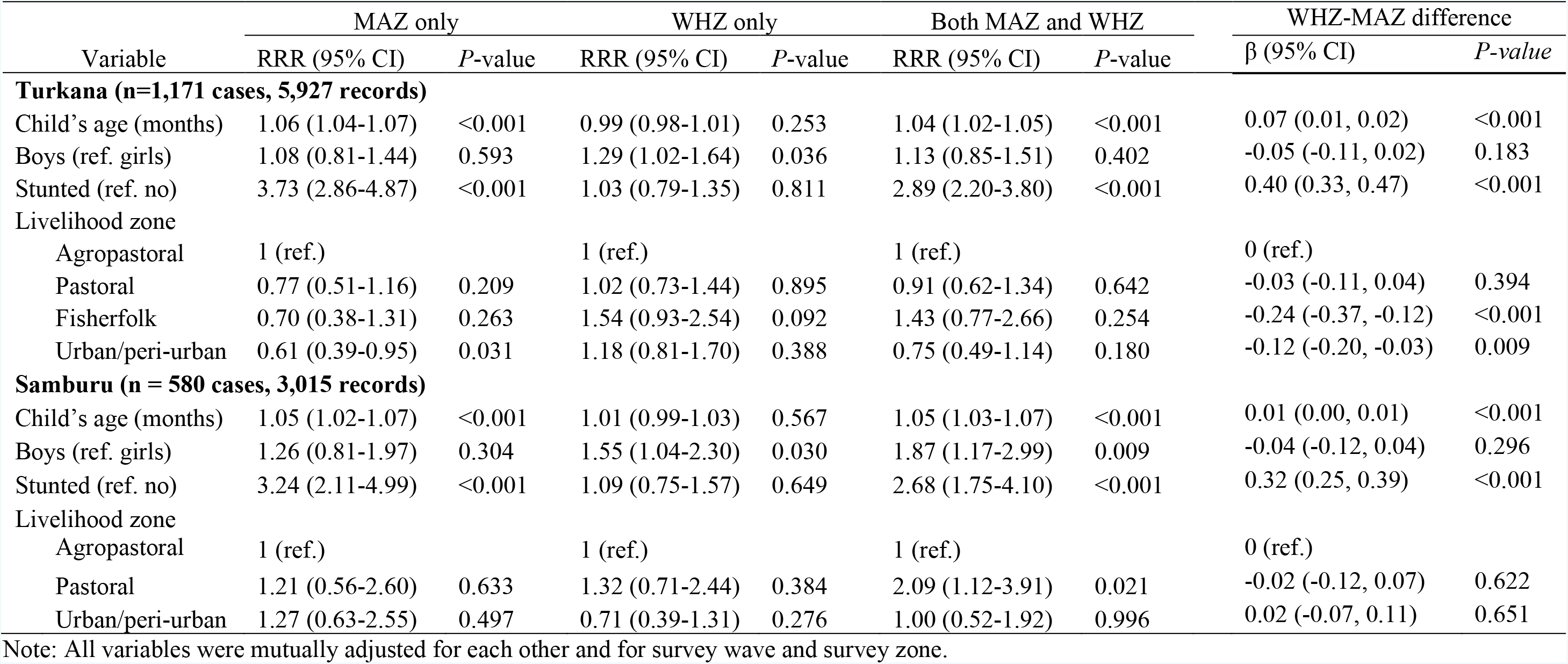
Factors associated with discordance and concordance between MAZ and WHZ in wasting diagnosis and the difference between WHZ and MAZ.

| Variable | MAZ only |  | WHZ only |  | Both MAZ and WHZ |  | WHZ-MAZ difference |  |
| --- | --- | --- | --- | --- | --- | --- | --- | --- |
| | RRR (95% CI) | *P*-value | RRR (95% CI) | *P*-value | RRR (95% CI) | *P*-value | $\beta$ (95% CI) | *P*-value |
| **Turkana (n=1,171 cases, 5,927 records)** |  |  |  |  |  |  |  |  |
| Child's age (months) | 1.06 (1.04-1.07) | <0.001 | 0.99 (0.98-1.01) | 0.253 | 1.04 (1.02-1.05) | <0.001 | 0.07 (0.01, 0.02) | <0.001 |
| Boys (ref. girls) | 1.08 (0.81-1.44) | 0.593 | 1.29 (1.02-1.64) | 0.036 | 1.13 (0.85-1.51) | 0.402 | -0.05 (-0.11, 0.02) | 0.183 |
| Stunted (ref. no) | 3.73 (2.86-4.87) | <0.001 | 1.03 (0.79-1.35) | 0.811 | 2.89 (2.20-3.80) | <0.001 | 0.40 (0.33, 0.47) | <0.001 |
| Livelihood zone |  |  |  |  |  |  |  |  |
| Agropastoral | 1 (ref.) |  | 1 (ref.) |  | 1 (ref.) |  | 0 (ref.) |  |
| Pastoral | 0.77 (0.51-1.16) | 0.209 | 1.02 (0.73-1.44) | 0.895 | 0.91 (0.62-1.34) | 0.642 | -0.03 (-0.11, 0.04) | 0.394 |
| Fisherfolk | 0.70 (0.38-1.31) | 0.263 | 1.54 (0.93-2.54) | 0.092 | 1.43 (0.77-2.66) | 0.254 | -0.24 (-0.37, -0.12) | <0.001 |
| Urban/peri-urban | 0.61 (0.39-0.95) | 0.031 | 1.18 (0.81-1.70) | 0.388 | 0.75 (0.49-1.14) | 0.180 | -0.12 (-0.20, -0.03) | 0.009 |
| **Samburu (n = 580 cases, 3,015 records)** |  |  |  |  |  |  |  |  |
| Child's age (months) | 1.05 (1.02-1.07) | <0.001 | 1.01 (0.99-1.03) | 0.567 | 1.05 (1.03-1.07) | <0.001 | 0.01 (0.00, 0.01) | <0.001 |
| Boys (ref. girls) | 1.26 (0.81-1.97) | 0.304 | 1.55 (1.04-2.30) | 0.030 | 1.87 (1.17-2.99) | 0.009 | -0.04 (-0.12, 0.04) | 0.296 |
| Stunted (ref. no) | 3.24 (2.11-4.99) | <0.001 | 1.09 (0.75-1.57) | 0.649 | 2.68 (1.75-4.10) | <0.001 | 0.32 (0.25, 0.39) | <0.001 |
| Livelihood zone |  |  |  |  |  |  |  |  |
| Agropastoral | 1 (ref.) |  | 1 (ref.) |  | 1 (ref.) |  | 0 (ref.) |  |
| Pastoral | 1.21 (0.56-2.60) | 0.633 | 1.32 (0.71-2.44) | 0.384 | 2.09 (1.12-3.91) | 0.021 | -0.02 (-0.12, 0.07) | 0.622 |
| Urban/peri-urban | 1.27 (0.63-2.55) | 0.497 | 0.71 (0.39-1.31) | 0.276 | 1.00 (0.52-1.92) | 0.996 | 0.02 (-0.07, 0.11) | 0.651 |

Note: All variables were mutually adjusted for each other and for survey wave and survey zone.

## Discussion

This 24-month longitudinal study shows that wasting prevalence measured using MUAC was consistently far lower than that measured using WHZ and MAZ in both Turkana and Samburu. Moreover, MUAC showed a dramatic decrease in wasting prevalence between Waves 1 and 6 while WHZ showed temporal fluctuations with no overall change between these survey time points in both counties. Wasting based on MAZ followed a pattern like that of WHZ in Turkana and Samburu, albeit with differences in the peak periods and an overall increase in Samburu. Our findings are consistent with other studies that have reported lower wasting prevalence based on MUAC than WHZ and MAZ in humanitarian settings (Bilukha & Leidman, 2018; Custodio et al., 2018). However, not all studies have found MUAC to underestimate the prevalence of wasting. An analysis of survey data from 47 countries found that the discrepancy in wasting diagnosis by WHZ and MUAC varied dramatically between countries with some having most children diagnosed as wasted by MUAC and others having almost all the children diagnosed as wasted by WHZ (Grellety & Golden, 2016). A cross-sectional study in Mozambique reported higher wasting prevalence based on MUAC than WHZ in 58.7% (27/46) of the districts assessed (Zaba et al., 2020). However, the prevalence of wasting (based on both WHZ and MUAC) in the Mozambican districts was generally lower (ranging from 0.0% to 10.9%) than in the studies in humanitarian settings similar to where our study was conducted. Higher wasting estimates based on WHZ and MAZ in Turkana and Samburu mean that these indicators can identify more children who are eligible for rehabilitation programs. The lower wasting prevalence based on MUAC means reliance on this indicator alone, as is the case in nutrition surveillance and “family MUAC” programs (UNICEF, Undated), can underestimate the burden of wasting in this and similar settings where color-coded MUAC tapes are widely used for rapid wasting screening.

Assessment of the sensitivity of MUAC and MAZ with reference to WHZ in wasting diagnosis showed that MAZ was more sensitive compared to MUAC, and the sensitivity of MAZ increased across the waves while that of MUAC decreased. WHZ had a better agreement with MAZ than MUAC. Because children aged at each subsequent wave, the decreasing sensitivity and agreement between MUAC and WHZ between Waves 1 and 6 suggests that MUAC is age dependent. In both counties, at Wave 1 when all children were younger than 3 years, the sensitivity of MUAC was just above 30% while that of MAZ was almost 40%. At Wave 6 when all the children were older than 2 years, the sensitivity of MUAC was below 10% while that of MAZ was above 50%. Thus, MUAC yielded more false negatives compared to MAZ, especially among older children. Overall, both MUAC and MAZ had high specificity, which is consistent with other studies (Lamsal et al., 2021). This is not surprising given that sensitivity and specificity have an inverse relationship. The PPV of MUAC ranged from 51% to 83% while NPV ranged from 78% to 91%. MAZ had predictive values similar to those of MUAC. Predictive values depend on the test’s sensitivity, specificity and the prevalence of the condition being screened (Celentano & Szklo, 2019). This means predictive values can vary by context and should be interpreted with this consideration. Taken together, our results suggest that with reference to WHZ, MAZ is a more reliable and valid indicator than MUAC in identifying wasted children.

Our study shows that classifications of wasting based on WHZ, MAZ, and MUAC are highly age, sex, and stunting status dependent. Considering MUAC and WHZ, older children were less likely to be classified as being wasted by MUAC alone or by both MUAC and WHZ but were more likely to be classified as wasted by WHZ alone. Compared to girls, boys were less likely to be classified as wasted by MUAC alone, but they were more likely to be classified as wasted by WHZ alone. In contrast, considering MAZ and WHZ, older children were more likely to be classified as wasted based on MAZ alone or both MAZ and WHZ. Boys were more likely to be classified as wasted by WHZ alone. Use of MUAC alone assumes that it does not change with age, whereas this study shows a clear increase in mean MUAC as children aged. Sex differences likely reflect different patterns of increase in MUAC in boys versus girls: girls have a larger increase in MUAC than boys. Stunted children were more likely to be classified as wasted by MUAC alone, MAZ alone, both MUAC and WHZ, and both MAZ and WHZ but not by WHZ alone. Our further analysis found no consistent differences in change in MUAC in stunted versus non-stunted children. However, stunted children had, on average, about 0.6 cm lower MUAC at different ages and waves, which can partly explain the difference.

Previous studies have used binary logistic regression to assess factors associated with wasting diagnosis based on WHZ, MUAC and MAZ as independent binary outcomes (Custodio et al., 2018) or linear regression models to ecologically assess factors associated with wasting prevalence based on MAZ and MUAC (Custodio et al., 2022). In our analysis, we used a different approach by treating classification of wasting by WHZ and MUAC and by WHZ and MAZ as polychotomous outcomes. Thus, we were able to assess the determinants of concordant and discordant classification of wasting based on the pairs of these indicators and provide new insights. Although our study and the studies mentioned above are not directly comparable given the differences in the analytic approaches, age, sex, and stunting status of the children play a role in their likelihood of being diagnosed as wasted depending on the indicator used. This means these factors should be accounted for in determining the thresholds for defining wasting based on MUAC. Stunted children were more likely to be identified by MUAC-based indicators as wasted because MUAC is a measure of muscle and fat mass, which is reduced in both wasted and stunted children (Briend, Khara, & Dolan, 2015; Villar et al., 2017).

The strengths of this study include a high follow-up rate, a representative sample, which allows for generalizability, and use of a longitudinal design, which allowed us to examine the performance of these indicators in wasting diagnosis over time in the same children as they aged. Our multivariable models used multiple observations per child and accounted for correlated measures.

In conclusion, the present study shows that MUAC is not age independent as implicitly assumed in the use of a single MUAC cut point in wasting diagnosis. Different classifications of wasting based on WHZ, MAZ, and MUAC are highly influenced by child’s age, sex, and stunting status. Our findings imply that MUAC alone is not appropriate for identifying wasted children, especially if they are stunted. MAZ is a more reliable and valid indicator compared to MUAC, but MAZ charts are harder to use than single MUAC cut points. Further research is needed to develop and test age and sex specific color-coded MUAC tapes for use in wasting screening in nutritional surveillance and community-based nutrition programs.

## Data Availability

The data underlying this study will be made available to researchers through the APHRC Microdata Portal.

https://aphrc.org/microdata-portal/

## Data sharing

In line with APHRC’s data access and sharing guidelines, the data underlying this study will be made available to researchers through the APHRC’s Microdata Portal (https://aphrc.org/microdata-portal/).

## Author contributions

CW, BM, and LA analyzed the data. FT, DA, VLF, CKL, AW and ES designed the study. CW, DA, BM, EA, HO, SE, and GC implemented fieldwork. CW, ES, FT, and EK-M provided study oversight. CW prepared the first draft of the manuscript and incorporated comments from the co-authors. All authors contributed to the revision of the manuscript and approved the final version.

