## Supplemental Tables and Figures for "Comparison of indicators for assessing wasting among children younger than 5 years: a longitudinal study in northern Kenya"

**Supplemental Table 1: Timing of data collection, expected season, actual rainfall received and early warning classification, Turkana**

| Data collection | | Actual rainfall received (mm) | Early warning classification † | | |
|---|---|---|---|---|---|
| **Timing** | **Expected season** | | **Pastoral** | **Agro-pastoral** | **Fisher folk** |
| Wave 1, May 2021 | Long rains | 46 | Alert | Normal | Normal |
| Wave 1, Jun 2021 | Long rains | 5 | Alert | Normal | Normal |
| Wave 2, Dec 2021 | Short rains | 6 | Alarm | Alarm | Alarm |
| Wave 3, Apr 2022 | Long rains | 18 | Alarm | Alert | Alarm |
| Wave 4, Sep 2022 | Short rains | 8 | Alarm | Alarm | Alarm |
| Wave 5, Feb 2023 | Dry season | 3 | Emergency | Emergency | Emergency |
| Wave 5, Mar 2023 | Dry season | 60 | Emergency | Emergency | Emergency |
| Wave 6, Aug 2023 | Cold lean season | 11 | Alert | Normal | Alert |

Source: Information on rainfall and early warning classification is from the National Drought Management Authority, Kenya

†The drought status is categorized into five levels: Normal, Alert, Alarm, Emergency, and Recovery.

**Supplemental Table 2: Timing of data collection, expected season, actual rainfall received and early warning classification, Samburu**

| Data collection | | Actual rainfall received (mm) | Early warning classification † | | |
|---|---|---|---|---|---|
| **Timing** | **Expected season** | | **Agropastoral** | **Pastoral north** | **Pastoral east** |
| Wave 1, Jun 2021 | Continental rains | 10 | Normal | Normal | Alarm |
| Wave 1, Jul 2021 | Continental rains | 12 | Normal | Normal | Alarm |
| Wave 2, Dec 2021 | Short rains | 45 | Alert | Alert | Alarm |
| Wave 3, Apr 2022 | Long rains | 99 | Alarm | Alarm | Alarm |
| Wave 4, Sep 2022 | Short rains | 8 | Alarm | Alarm | Alarm |
| Wave 5, Feb 2023 | Dry season | 5 | Alarm | Alarm | Alarm |
| Wave 5, Mar 2023 | Dry season | missing | Alarm | Alarm | Alarm |
| Wave 6, Aug 2023 | Continental rains | 26-50 | Alert | Alert | Alert |

Source: Information on rainfall and early warning classification is from the National Drought Management Authority in Kenya
†The drought status is categorized into five levels: Normal, Alert, Alarm, Emergency, and Recovery.

Supplemental Table 3: Characteristics of study participants

| | Turkana | | | | | | Samburu | | | | | |
|---|---|---|---|---|---|---|---|---|---|---|---|---|
| | Wave 1 (N=1,211) | Wave 2 (N=1,068) | Wave 3 (N=1,019) | Wave 4 (N=1,134) | Wave 5 (N=1,082) | Wave 6 (N=1,156) | Wave 1 (N=586) | Wave 2 (N=556) | Wave 3 (N=522) | Wave 4 (N=549) | Wave 5 (N=543) | Wave 6 (N=544) |
| **Turkana** | | | | | | | | | | | | |
| Child's age (months), Mean ± SD | 16.8 ± 10.6 | 22.7± 10.6 | 28.5 ± 10.6 | 34.1 ± 10.5 | 38.4 ± 10.5 | 44.1 ± 10.5 | 14.4 ± 10.0 | 19.3 ± 10.0 | 24.1 ± 10.0 | 29.9 ± 10.0 | 35.2 ± 10.2 | 39.9 ± 10.1 |
| Child's sex | | | | | | | | | | | | |
| Girls | 45.2 | 47.3 | 46.9 | 46.2 | 48.0 | 47.2 | 47.8 | 48.4 | 48.9 | 47.7 | 48.1 | 48.7 |
| Boys | 54.8 | 52.7 | 53.1 | 53.8 | 52.0 | 52.8 | 52.2 | 51.6 | 51.1 | 52.3 | 51.9 | 51.3 |
| Stunted | 25.9 | 27.8 | 27.9 | 30.4 | 29.3 | 26.5 | 24.5 | 35.9 | 37.2 | 41.4 | 43.6 | 41.3 |
| Survey zone | | | | | | | | | | | | |
| Central | 21.0 | 20.6 | 19.8 | 20.3 | 21.6 | 21.4 | 7.1 | 6.8 | 7.0 | 6.7 | 6.7 | 6.7 |
| North | 25.6 | 26.6 | 28.6 | 26.2 | 27.2 | 26.4 | 77.1 | 77.9 | 78.0 | 78.1 | 77.5 | 77.7 |
| South | 24.1 | 24.8 | 21.7 | 24.0 | 22.4 | 23.9 | -- | -- | -- | -- | -- | -- |
| West | 29.2 | 28.0 | 30.0 | 29.5 | 28.8 | 28.3 | -- | -- | -- | -- | -- | -- |
| East | -- | -- | -- | -- | -- | -- | 15.8 | 15.3 | 14.9 | 15.2 | 15.8 | 15.5 |
| Livelihood zone | | | | | | | | | | | | |
| Agropastoral | 14.8 | 15.5 | 16.5 | 10.1 | 8.6 | 10.4 | 5.2 | 6.0 | 5.5 | 4.9 | 4.9 | 4.9 |
| Pastoral | 63.9 | 44.8 | 58.7 | 67.5 | 68.6 | 67.2 | 83.4 | 80.7 | 86.0 | 80.2 | 81.6 | 81.5 |
| Urban/peri-urban | 6.5 | 21.5 | 6.5 | 8.7 | 8.1 | 8.0 | 11.4 | 13.3 | 8.5 | 14.9 | 13.5 | 13.6 |
| Fisherfolk | 14.8 | 18.2 | 18.2 | 13.7 | 14.7 | 14.4 | -- | -- | -- | -- | -- | -- |

Note: The numbers are percentages except for age.

Supplemental Table 4: Mean WHZ, MUAC and MAZ by county, survey wave, and sex

| | Turkana (mean ± SD) | | | Samburu (mean ± SD) | | |
|---|---|---|---|---|---|---|
| | WHZ | MUAC | MAZ | WHZ | MUAC | MAZ |
| Wave 1 | | | | | | |
| Girls | -1.08 ± 1.48 | 13.52 ± 1.10 | -0.92 ± 1.12 | -1.26 ± 1.60 | 13.56 ± 1.16 | -0.79 ± 0.95 |
| Boys | -1.08 ± 1.46 | 13.66 ± 1.07 | -1.16 ± 1.11 | -1.27 ± 1.21 | 13.91 ± 0.99 | -0.85 ± 1.05 |
| Overall | -1.08 ± 1.47 | 13.59 ± 1.09 | -1.05 ± 1.12 | -1.26 ± 1.43 | 13.73 ± 1.10 | -0.82 ± 1.00 |
| N | 1,117 | 919 | 889 | 533 | 459 | 459 |
| Wave 2 | | | | | | |
| Girls | -1.08 ± 1.02 | 13.77 ± 1.02 | -0.95 ± 0.99 | -0.79 ± 1.14 | 13.62 ± 0.96 | -0.93 ± 0.85 |
| Boys | -1.24 ± 1.06 | 13.86 ± 1.06 | -1.20 ± 1.08 | -1.24 ± 1.08 | 13.71 ± 0.86 | -1.24 ± 0.91 |
| Overall | -1.16 ± 1.04 | 13.82 ± 1.04 | -1.08 ± 1.05 | -1.01 ± 1.13 | 13.66 ± 0.92 | -1.08 ± 0.89 |
| N | 1,046 | 1,009 | 1,008 | 517 | 530 | 528 |
| Wave 3 | | | | | | |
| Girls | -1.33 ± 0.99 | 13.69 ± 0.97 | -1.27 ± 0.98 | -0.96 ± 0.96 | 13.77 ± 0.94 | -1.00 ± 0.85 |
| Boys | -1.54 ± 1.01 | 13.74 ± 1.05 | -1.46 ± 1.04 | -1.39 ± 0.93 | 13.80 ± 0.91 | -1.29 ± 0.92 |
| Overall | -1.44 ± 1.00 | 13.72 ± 1.01 | -1.37 ± 1.01 | -1.17 ± 0.97 | 13.78 ± 0.92 | -1.14 ± 0.90 |
| N | 979 | 964 | 959 | 489 | 506 | 506 |
| Wave 4 | | | | | | |
| Girls | -1.28 ± 0.99 | 13.88 ± 1.00 | -1.34 ± 0.95 | -1.22 ± 0.99 | 14.08 ± 1.04 | -0.96 ± 0.97 |
| Boys | -1.35 ± 0.97 | 14.01 ± 1.08 | -1.43 ± 1.07 | -1.60 ± 0.92 | 14.01 ± 0.96 | -1.29 ± 0.95 |
| Overall | -1.32 ± 0.98 | 13.95 ± 1.04 | -1.39 ± 1.02 | -1.40 ± 0.97 | 14.05 ± 1.00 | -1.12 ± 0.97 |
| N | 1,079 | 1,096 | 1,082 | 547 | 545 | 545 |
| Wave 5 | | | | | | |
| Girls | -1.34 ± 0.95 | 14.02 ± 1.03 | -1.38 ± 0.95 | -1.23 ± 0.81 | 14.11 ± 0.96 | -1.16 ± 0.88 |
| Boys | -1.42 ± 0.98 | 14.06 ± 1.03 | -1.50 ± 1.01 | -1.64 ± 0.88 | 13.92 ± 0.97 | -1.55 ± 0.96 |
| Overall | -1.38 ± 0.96 | 14.04 ± 1.03 | -1.44 ± 0.98 | -1.43 ± 0.87 | 14.02 ± 0.97 | -1.35 ± 0.94 |
| N | 1,035 | 1,070 | 1,070 | 543 | 543 | 543 |
| Wave 6 | | | | | | |
| Girls | -1.25 ± 0.99 | 14.46 ± 1.03 | -1.17 ± 0.90 | -1.11 ± 1.00 | 14.30 ± 0.94 | -1.18 ± 0.86 |
| Boys | -1.33 ± 0.97 | 14.40 ± 1.03 | -1.30 ± 0.94 | -1.45 ± 1.03 | 14.19 ± 1.00 | -1.42 ± 0.98 |
| Overall | -1.29 ± 0.98 | 14.43 ± 1.03 | -1.24 ± 0.92 | -1.27 ±1.03 | 14.25 ± 0.97 | -1.30 ± 0.93 |
| N | 1,026 | 1,125 | 1,012 | 529 | 540 | 529 |

Supplemental Table 5: Prevalence of wasting based on different anthropometric indicators stratified by child's sex

| | Turkana | | | Samburu | | |
|---|---|---|---|---|---|---|
| | WHZ (%) | MUAC (%) | MAZ (%) | WHZ (%) | MUAC (%) | MAZ (%) |
| Wave 1 | | | | | | |
| Girls | 23.1 | 15.6 | 18.1 | 23.7 | 14.5 | 9.8 |
| Boys | 20.6 | 11.7 | 20.5 | 22.8 | 10.1 | 19.1 |
| N | 1,117 | 919 | 889 | 533 | 459 | 459 |
| Wave 2 | | | | | | |
| Girls | 18.6 | 10.4 | 14.2 | 10.0 | 15.7 | 7.8 |
| Boys | 21.3 | 10.5 | 19.8 | 23.9 | 8.6 | 21.3 |
| N | 1,046 | 1,009 | 1,008 | 517 | 530 | 528 |
| Wave 3 | | | | | | |
| Girls | 22.8 | 11.2 | 23.3 | 11.7 | 9.0 | 11.3 |
| Boys | 33.4 | 13.8 | 28.9 | 32.0 | 5.9 | 22.4 |
| N | 979 | 964 | 959 | 489 | 506 | 506 |
| Wave 4 | | | | | | |
| Girls | 22.0 | 8.0 | 26.1 | 26.4 | 3.1 | 14.2 |
| Boys | 21.8 | 8.5 | 26.7 | 35.1 | 2.8 | 26.2 |
| N | 1,079 | 1,096 | 1,082 | 547 | 545 | 545 |
| Wave 5 | | | | | | |
| Girls | 24.1 | 5.0 | 29.0 | 15.1 | 4.6 | 20.3 |
| Boys | 25.8 | 8.5 | 33.3 | 40.7 | 6.3 | 33.2 |
| N | 1,035 | 1,070 | 1,070 | 543 | 543 | 543 |
| Wave 6 | | | | | | |
| Girls | 18.4 | 3.3 | 16.0 | 17.3 | 2.4 | 14.8 |
| Boys | 25.5 | 1.8 | 25.0 | 30.8 | 2.3 | 27.9 |
| N | 1,026 | 1,125 | 1,012 | 529 | 540 | 529 |

Supplemental Figure 1: Seasonality of wasting based on different anthropometric indicators in Turkana and Samburu

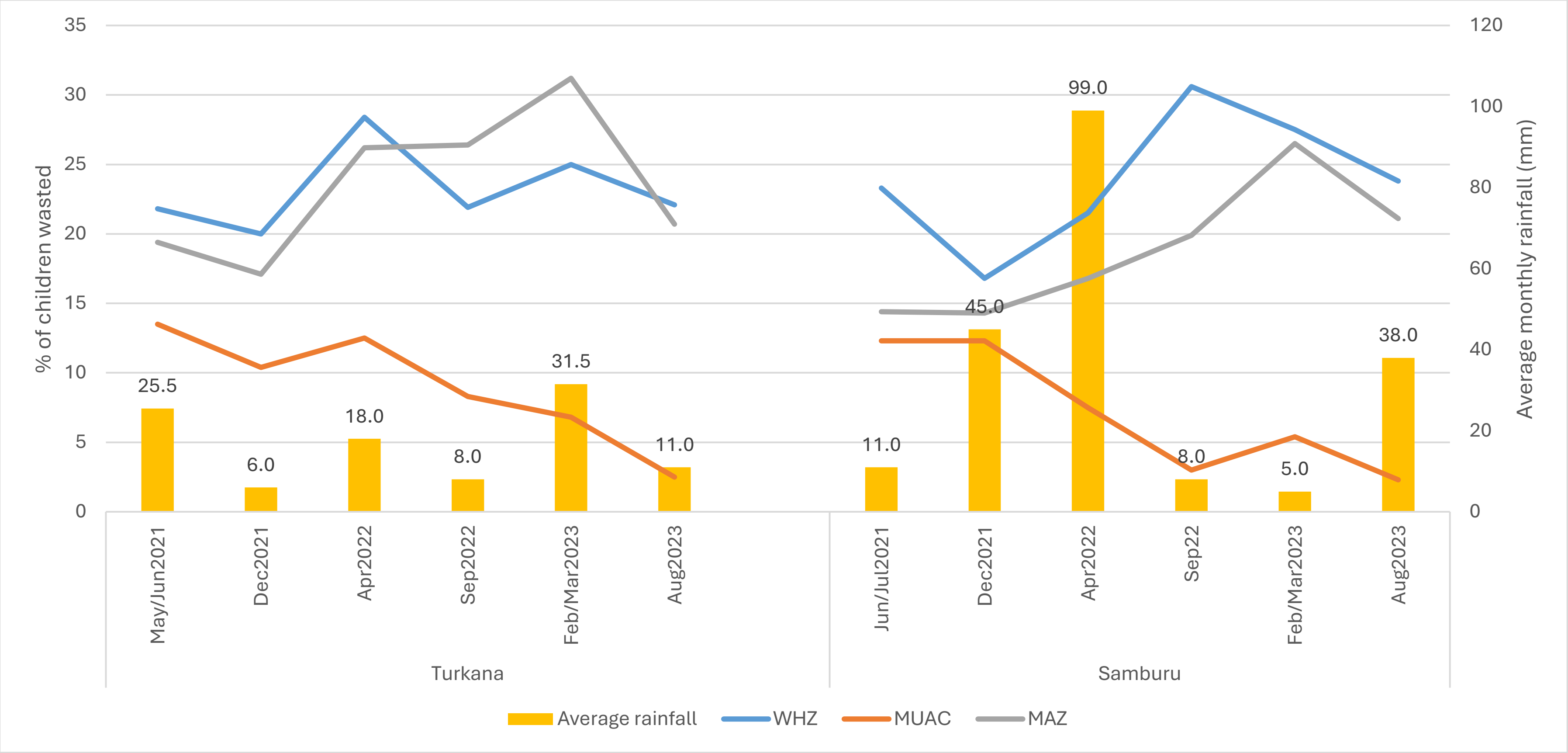

Supplemental Table 6: Correlation of WHZ with MUAC and MAZ by county and survey wave

| | Pearson's correlation coefficient (rho) | | | |
|---|---|---|---|---|
| | Turkana | | Samburu | |
| | WHZ vs. MUAC | WHZ vs. MAZ | WHZ vs. MUAC | WHZ vs. MAZ |
| Wave 1 | 0.472 | 0.496 | 0.656 | 0.695 |
| Wave 2 | 0.667 | 0.691 | 0.651 | 0.729 |
| Wave 3 | 0.692 | 0.698 | 0.727 | 0.782 |
| Wave 4 | 0.681 | 0.690 | 0.736 | 0.797 |
| Wave 5 | 0.668 | 0.684 | 0.735 | 0.796 |
| Wave 6 | 0.615 | 0.647 | 0.679 | 0.721 |

Supplemental Table 7: Mean of the difference between WHZ and MAZ by wave and child's age

| | Turkana | | Samburu | |
|---|---|---|---|---|
| | N | Mean ± SD | N | Mean ± SD |
| Wave 1 | | | | |
| ≤24 months | 688 | -0.16 ±1.31 | 364 | -0.31 ± 0.84 |
| >24 months | 278 | 0.15 ± 1.09 | 111 | -0.08 ± 0.69 |
| Total | 966 | -0.06 ± 1.25 | 475 | -0.26 ± 0.82 |
| Wave 2 | | | | |
| ≤24 months | 565 | -0.30 ± 0.79 | 325 | -0.04 ± 0.77 |
| >24 months | 443 | 0.14 ± 0.75 | 168 | 0.04 ± 0.78 |
| Total | 1,008 | -0.09 ±0.80 | 493 | -0.01 ± 0.80 |
| Wave 3 | | | | |
| ≤24 months | 382 | -0.40 ± 0.69 | 239 | -0.23 ± 0.66 |
| >24 months | 558 | 0.19 ± 0.74 | 239 | 0.16 ± 0.58 |
| Total | 940 | -0.06 ± 0.78 | 478 | -0.05 ± 0.70 |
| Wave 4 | | | | |
| ≤24 months | 247 | -0.34 ± 0.73 | 187 | -0.50 ± 0.62 |
| >24 months | 821 | 0.16 ± 0.70 | 358 | -0.15 ± 0.62 |
| Total | 1,068 | 0.06 ± 0.74 | 545 | -0.28 ± 0.06 |
| Wave 5 | | | | |
| ≤24 months | 106 | -0.38 ± 0.77 | 73 | -0.36 ± 0.48 |
| >24 months | 929 | 0.11 ± 0.74 | 470 | -0.02 ± 0.61 |
| Total | 1,035 | 0.07 ± 0.76 | 543 | -0.08 ± 0.60 |
| Wave 6 | | | | |
| ≤24 months | 0 | -- | 0 | -- |
| >24 moths | 1,009 | -0.05 ± 0.75 | 529 | 0.02 ± 0.79 |
| Total | 6,026 | -0.02 ± 0.86 | 3,063 | -0.11 ± 0.72 |

Supplemental Figure 2: Sensitivity, Specificity, and predictive values of MUAC and MAZ with reference to WHZ, Turkana

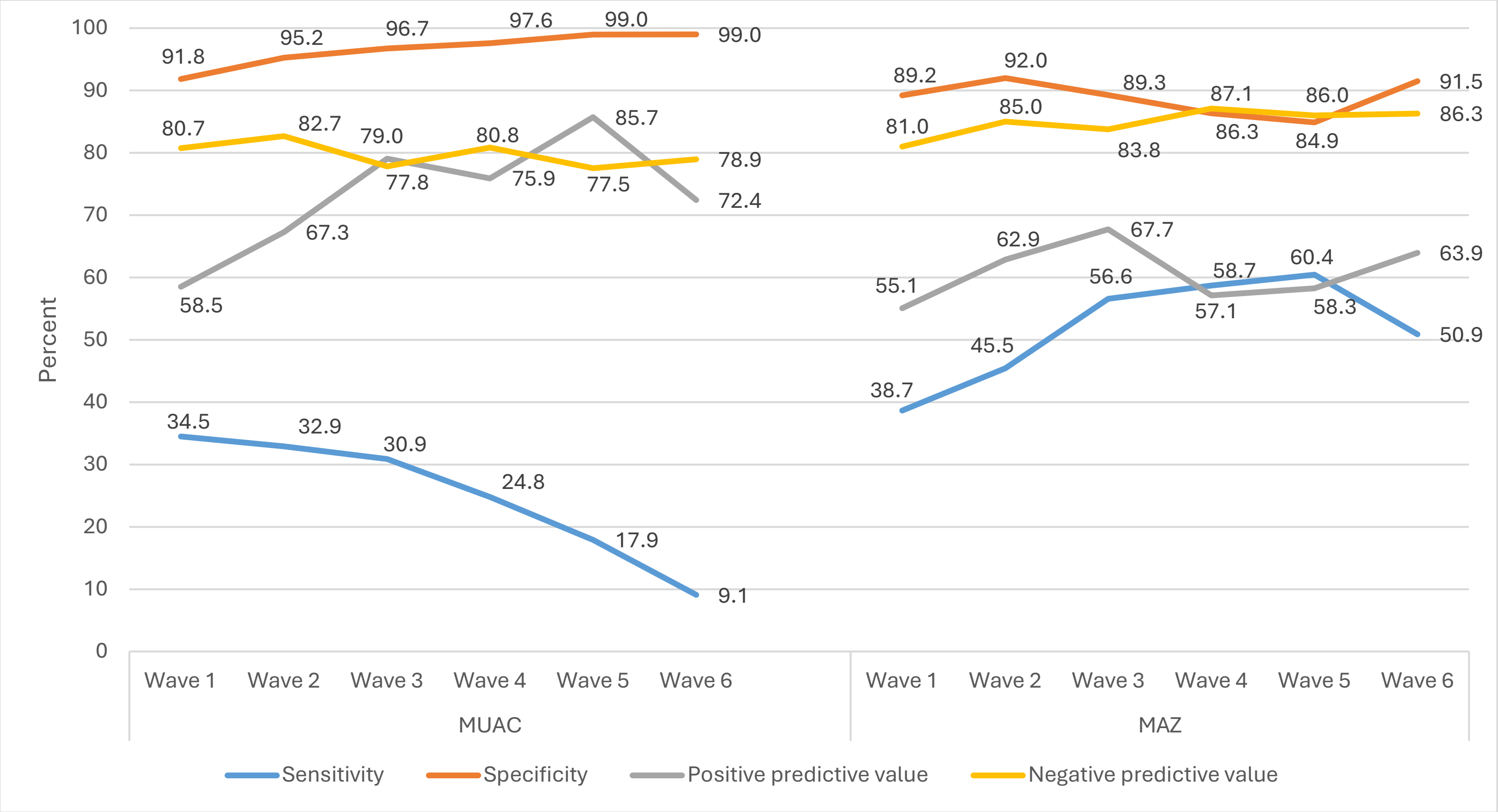

Supplemental Figure 3: Sensitivity, Specificity, and predictive values of MUAC and MAZ with reference to WHZ, Samburu

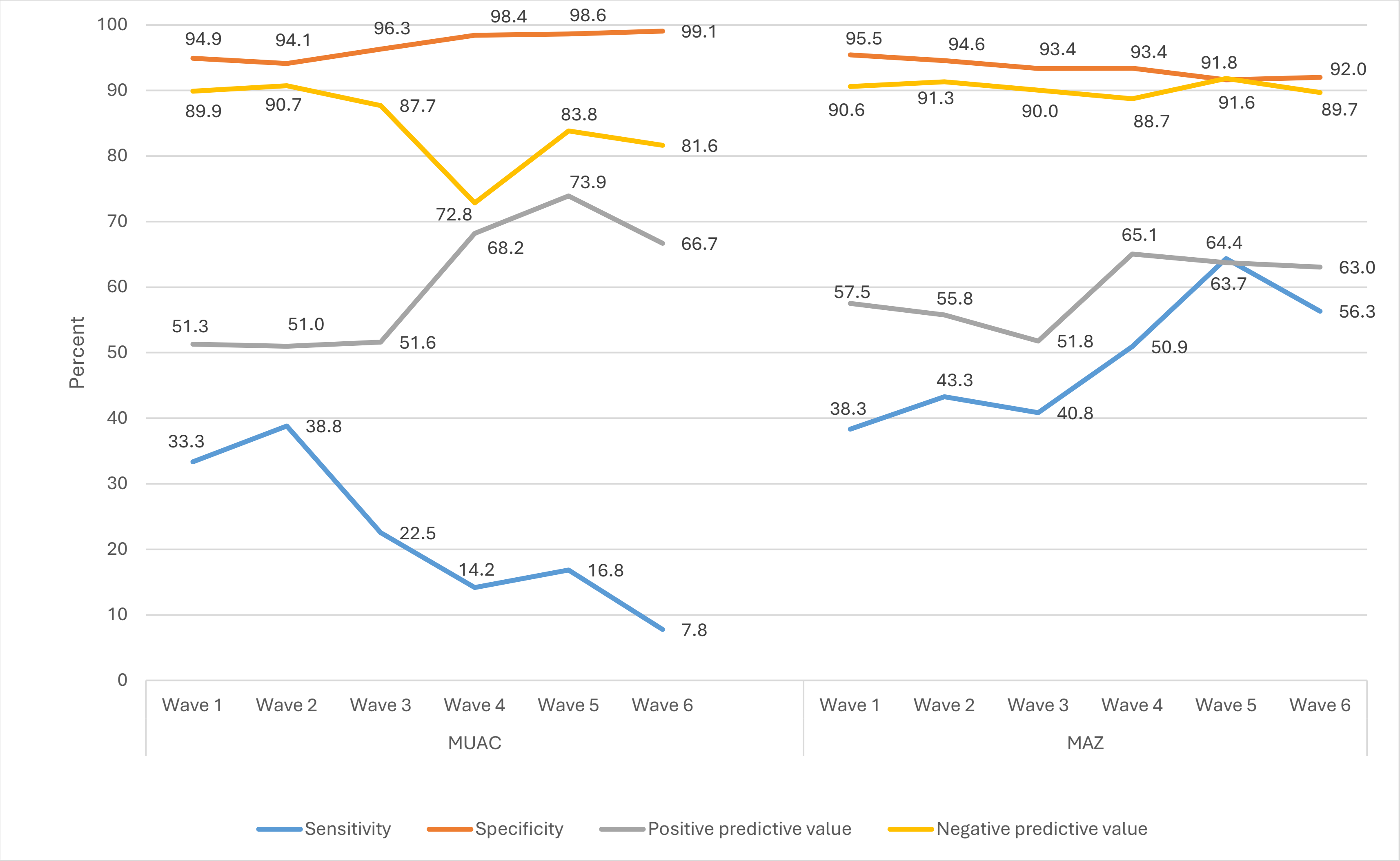

Supplemental Table 8: The percentage of children by wasting status based on a combination of different anthropometric indicators by survey wave and county

| | Wave 1 (%) | Wave 2 (%) | Wave 3 (%) | Wave 4 (%) | Wave 5 (%) | Wave 6 (%) | Total (%) |
|---|---|---|---|---|---|---|---|
| **Turkana** | | | | | | | |
| Wasting based on WHZ and MUAC | | | | | | | |
| Not wasted by WHZ and MUAC | 72.6 | 76.6 | 69.2 | 76.2 | 74.1 | 77.1 | 73.4 |
| Wasted by MUAC only | 5.2 | 3.4 | 2.3 | 1.9 | 1.0 | 0.8 | 2.4 |
| Wasted by WHZ only | 14.0 | 13.0 | 18.4 | 15.8 | 19.2 | 20.4 | 16.8 |
| Wasted by WHZ and MUAC | 8.3 | 7.0 | 10.1 | 6.1 | 5.8 | 1.7 | 6.5 |
| N | 913 | 1,007 | 943 | 1,079 | 1,035 | 1,026 | 6,003 |
| Wasting based on WHZ and MAZ | | | | | | | |
| Not wasted by WHZ and MAZ | 68.4 | 72.8 | 62.6 | 65.8 | 61.2 | 70.1 | 66.7 |
| Wasted by MAZ only | 8.7 | 7.2 | 10.2 | 11.9 | 13.8 | 7.6 | 10.0 |
| Wasted by WHZ only | 12.3 | 10.1 | 11.2 | 7.9 | 7.1 | 9.2 | 9.5 |
| Wasted by WHZ and MAZ | 10.7 | 9.9 | 16.0 | 14.4 | 17.8 | 13.0 | 13.7 |
| N | 884 | 1,006 | 938 | 1,065 | 1,035 | 1,006 | 5,934 |
| **Samburu** | | | | | | | |
| Wasting based on WHZ and MUAC | | | | | | | |
| Not wasted by WHZ and MUAC | 77.2 | 77.0 | 75.5 | 68.8 | 71.8 | 75.4 | 74.1 |
| Wasted by MUAC only | 4.7 | 6.0 | 2.4 | 0.6 | 0.7 | 0.9 | 2.4 |
| Wasted by WHZ only | 10.7 | 10.7 | 16.8 | 28.2 | 22.8 | 22.3 | 19.1 |
| Wasted by WHZ and MUAC | 7.4 | 6.3 | 5.3 | 2.4 | 4.7 | 1.4 | 4.4 |
| N | 434 | 493 | 478 | 545 | 543 | 529 | 3,022 |
| Wasting based on WHZ and MAZ | | | | | | | |
| Not wasted by WHZ and MAZ | 75.1 | 76.4 | 70.2 | 63.9 | 64.9 | 68.1 | 69.5 |
| Wasted by MAZ only | 6.8 | 6.6 | 7.7 | 5.5 | 7.6 | 8.2 | 7.1 |
| Wasted by WHZ only | 10.1 | 9.2 | 12.6 | 16.2 | 8.5 | 10.8 | 11.3 |
| Wasted by WHZ and MAZ | 8.0 | 7.8 | 9.4 | 14.5 | 18.9 | 12.9 | 12.2 |
| N | 434 | 491 | 478 | 545 | 543 | 529 | 3,020 |
